# The distribution of paediatric forearm fractures. A five-year retrospective cohort study of 4,546 forearm fractures in children

**DOI:** 10.1101/2025.04.01.25325050

**Authors:** Hans-Christen Husum, Søren Kold, Ole Rahbek

## Abstract

**Background:** Forearm fractures are the most common fractures in children, accounting for 41% of all paediatric fractures. Most research focuses on distal forearm fractures, but studies encompassing the entire forearm are limited. This retrospective study describes the distribution and patterns of paediatric forearm fractures over a five-year period.

**Methods:** We conducted a retrospective cohort study of children aged 0-15 years who received a radiograph of the forearm, wrist or elbow between March 2019 and December 2023 in the study region. Fractures were manually identified and registered from radiological reports. Fracture location, type (complete/incomplete), and epiphyseal involvement were analyzed across different age groups. Statistical analysis was performed using chi-square tests and descriptive statistics.

**Results:** We identified 4,547 forearm fractures from 4,291 children. The median age was 10 years, and 57% of the patients were male. Fracture patterns varied significantly across age groups (p<0.001), with older children experiencing more distal, complete, radial, and epiphyseal fractures. Younger children had a higher proportion of incomplete fractures and fewer distal or epiphyseal fractures. No significant differences in Salter-Harris classifications were found between age groups (p=0.69).

**Conclusion:** Fracture patterns in paediatric forearm fractures vary with age, with older children showing a higher incidence of complete, distal, and epiphyseal fractures. This study provides a detailed characterization of paediatric forearm fractures, which may inform clinical management and preventive strategies, particularly in tailoring age-specific care. Further research should explore the long-term outcomes of these fracture patterns.

## Introduction

Forearm fractures are a common injury in children, making up 41% of all paediatric fractures, with the distal forearm being the most commonly affected ^1–3^ and adolescent boys having the highest incidence ^4^.

As such, there is an abundance of research focusing on paediatric distal radius fractures ^5–7^, but detailed studies of fracture patterns of the entire paediatric forearm are lacking.

While there are large-scale epidemiological studies conducted through patient registries in Scandinavia ^8,9^, these registries rely on the ICD-10 diagnostic code system, which does not feature the level of detail to distinguish between complete/incomplete fractures or epiphyseal fractures. Further, the validity of paediatric fracture diagnoses within some of the registries is unknown^10^. Despite the large amount of available scientific evidence, a recent systematic review of paediatric fracture incidences made by the European Society of Paediatric Radiology noted a lack of validated research databases in the available body of evidence ^11^.

In the North Denmark Region, it is standard practice for any acute paediatric radiograph to be described by an expert musculoskeletal radiographer/radiologist within 24-hours of acquisition.

In designing this study, we therefore chose to manually extract and register paediatric forearm fractures using written radiological reports as the validity and informative level regarding diagnostic codes of paediatric forearm fractures can be questioned

The purpose of this study was to describe the patterns of paediatric forearm fractures, using written radiological reports, and how these patterns might change across different age groups.

## Methods

### Study design

This was a retrospective cohort study of children diagnosed with an acute forearm fracture during any type of visit (emergency room/inpatient/outpatient) to any of the three institutions in the North Denmark Region in a five-year period between March 1^st^ 2019 to December 31^st^ 2023. Reporting follows the STROBE guidelines for reporting on observational studies ^12^

### Participants

All radiological reports of radiographs of the forearm, wrist or elbow of children aged 0-15 in the study period were screened for eligibility. Radiological reports were identified and extracted using examination codes (UXRF25/30/40) and examination date within the study period. To avoid duplicates or radiographs taken as part of routine follow-up, radiographs for the same patient and extremity had to be at least three months apart.

We included all radiological reports where a fracture was detected and excluded examinations where a written radiological report was missing.

### Variables

From the radiological reports we extracted the patients’ civil registration number which was used to register the patients’ gender, age at radiograph and to avoid duplicate examinations of the same patient. Further, from the radiological report of the radiographs, we extracted information of the fractures including location of the fracture, type of fracture (complete/incomplete/mixed), epiphyseal fractures and Salter Harris (SH) classification.

A fracture was registered when it was specifically stated in the radiological report. Suspected fractures or cases where the radiologist was in doubt was not registered as fractures. An epiphyseal fracture was registered when physeal involvement was described and the SH classification was registered when the SH type was explicitly stated in the radiological report.

The radiological reports were collected on the 14^th^ of May 2024. All data was manually extracted by four medical students under the direct instruction and supervision by the first author. Data was entered and registered directly in a GDPR compliant secure REDCap database hosted at Aalborg University Hospital, Aalborg, Denmark.

### Statistical methods

Descriptive analysis was performed, and results are presented in count and percentages, with age presented as median and interquartile interval. Proportions of anatomical fracture types, fracture localisations and SH types were calculated and compared across age groups using chi-square statistics. Normal distribution of data was inspected using quantile-quantile plots and a 5% significance level was applied. All calculations were performed using Stata version 18.0 (StataCorp LLC, College Station Texas, USA).

### Ethics and approvals

Upon query, approval from the Region North Denmark institutional ethical committee was deemed unnecessary (case no 2-1-02-1167-24). The study was registered in the institutional research database (case no F2024-043) and vicarious consent for data collection for the purposes of this study was obtained (case no 2024-010476).

## Results

During the study period, we identified 17,879 radiographs from 12,728 children. Of these, 2,700 radiographs (15%) did not have a radiological report. For the remaining 15,179 radiographs with reports, 4,547 radiographs, involving 4,291 children, included a description of a fracture (Fig 1).

**Figure 1:**
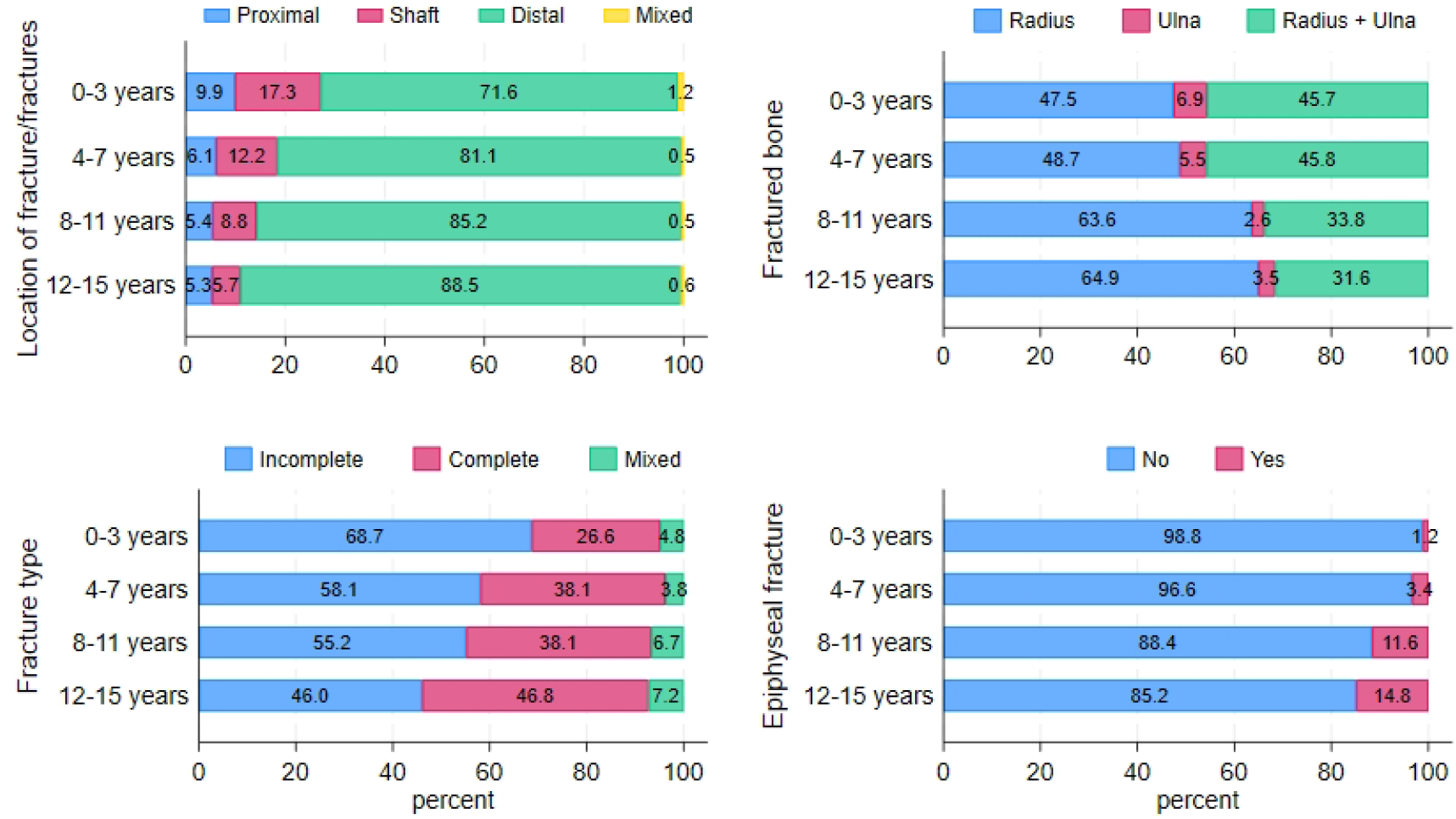
flowchart depicting the inclusion process.

The majority of the patients were male 2590 (57%) versus 1957 female (43%) and 501 patients had more than one forearm fracture during the five-year study period. The median age of the patient at fracture diagnosis was 10 years (IQI 6 to 12).

There was a significant difference in distribution across age groups for affected bones, fracture location, fracture type and epiphyseal fractures (p<0.001). We saw a tendency of fractures becoming proportionally more distal, complete, affecting the radius and involving the epiphysis as the children became older (Table 1, Figure 2).

**Table 1:**
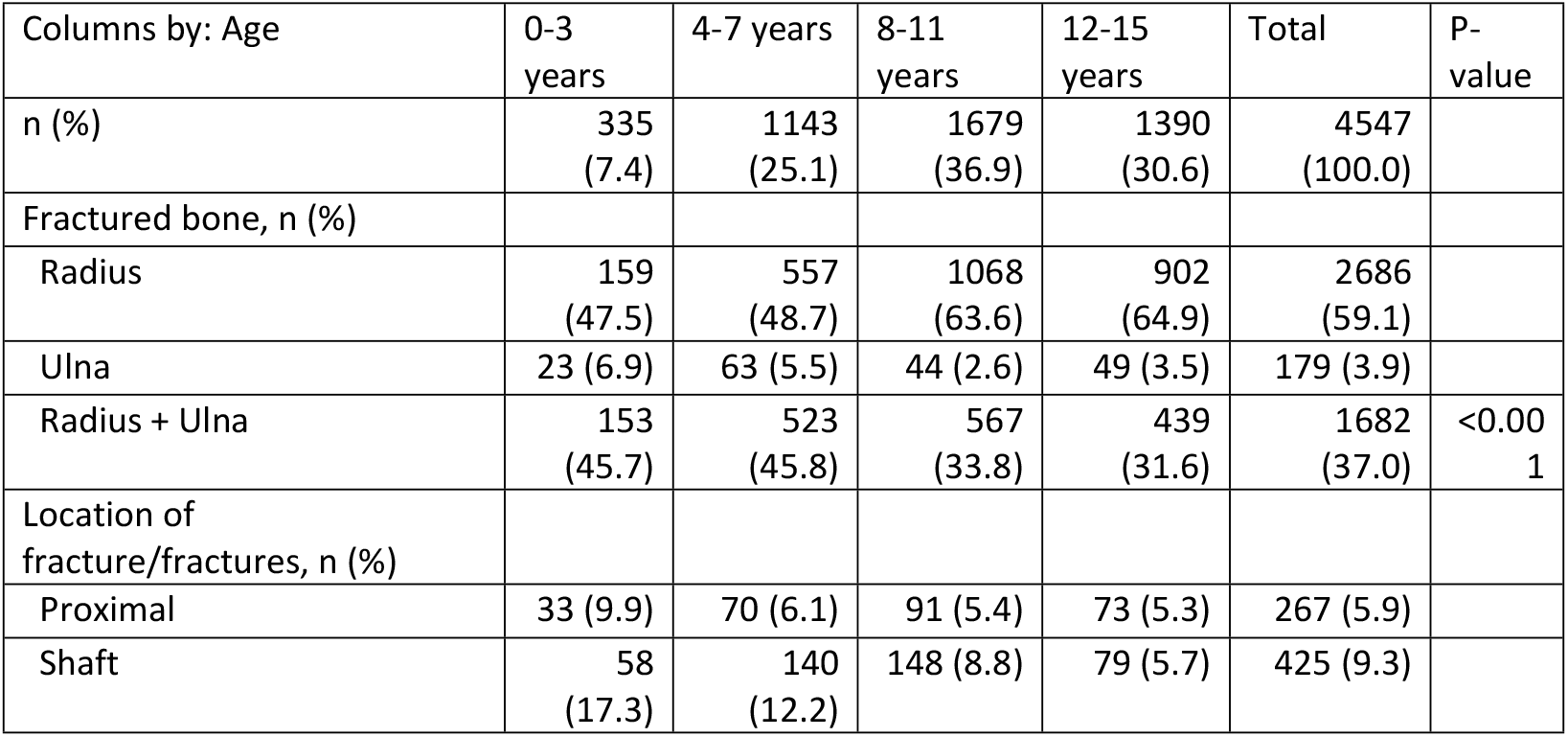
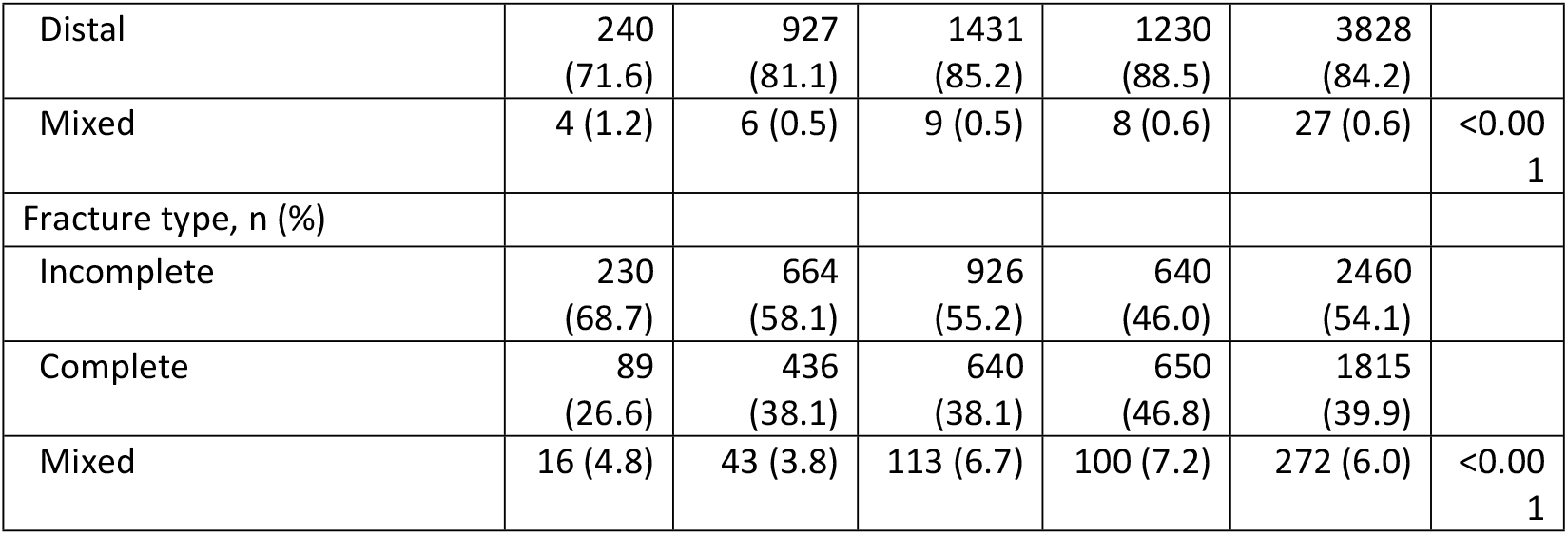
distribution of fracture locations stratified by age group.

**Table 2:** distribution of epiphyseal fracture and Salter Harris (SH) type stratified by age group.

| Table 2 | 0-3<br>years | 4-7<br>years | 8-11<br>years | 12-15<br>years | Total | P-value |
| --- | --- | --- | --- | --- | --- | --- |
| n (%) | 335<br>(7.4) | 1143<br>(25.1) | 1679<br>(36.9) | 1390<br>(30.6) | 4547<br>(100.0) |  |
| Epiphyseal fracture, n (%) |  |  |  |  |  |  |
| No | 331<br>(98.8) | 1104<br>(96.6) | 1485<br>(88.4) | 1183<br>(85.2) | 4103<br>(90.3) |  |
| Yes | 4 (1.2) | 39 (3.4) | 194<br>(11.6) | 206<br>(14.8) | 443 (9.7) | <0.001 |
| Type of epiphyseal fracture (Salter Harris), n (%) |  |  |  |  |  |  |
| SH1 | 0 (0.0) | 1 (2.6) | 10 (5.2) | 8 (3.9) | 19 (4.3) |  |
| SH2 | 4<br>(100.0) | 37<br>(94.9) | 156<br>(80.4) | 156<br>(75.7) | 353<br>(79.7) |  |
| SH3 | 0 (0.0) | 0 (0.0) | 5 (2.6) | 10 (4.9) | 15 (3.4) |  |
| SH4 | 0 (0.0) | 0 (0.0) | 3 (1.5) | 3 (1.5) | 6 (1.4) |  |
| SH5 | 0 (0.0) | 0 (0.0) | 0 (0.0) | 1 (0.5) | 1 (0.2) |  |
| Not described | 0 (0.0) | 1 (2.6) | 20<br>(10.3) | 28<br>(13.6) | 49 (11.1) | 0.69 |

**Figure 2:**
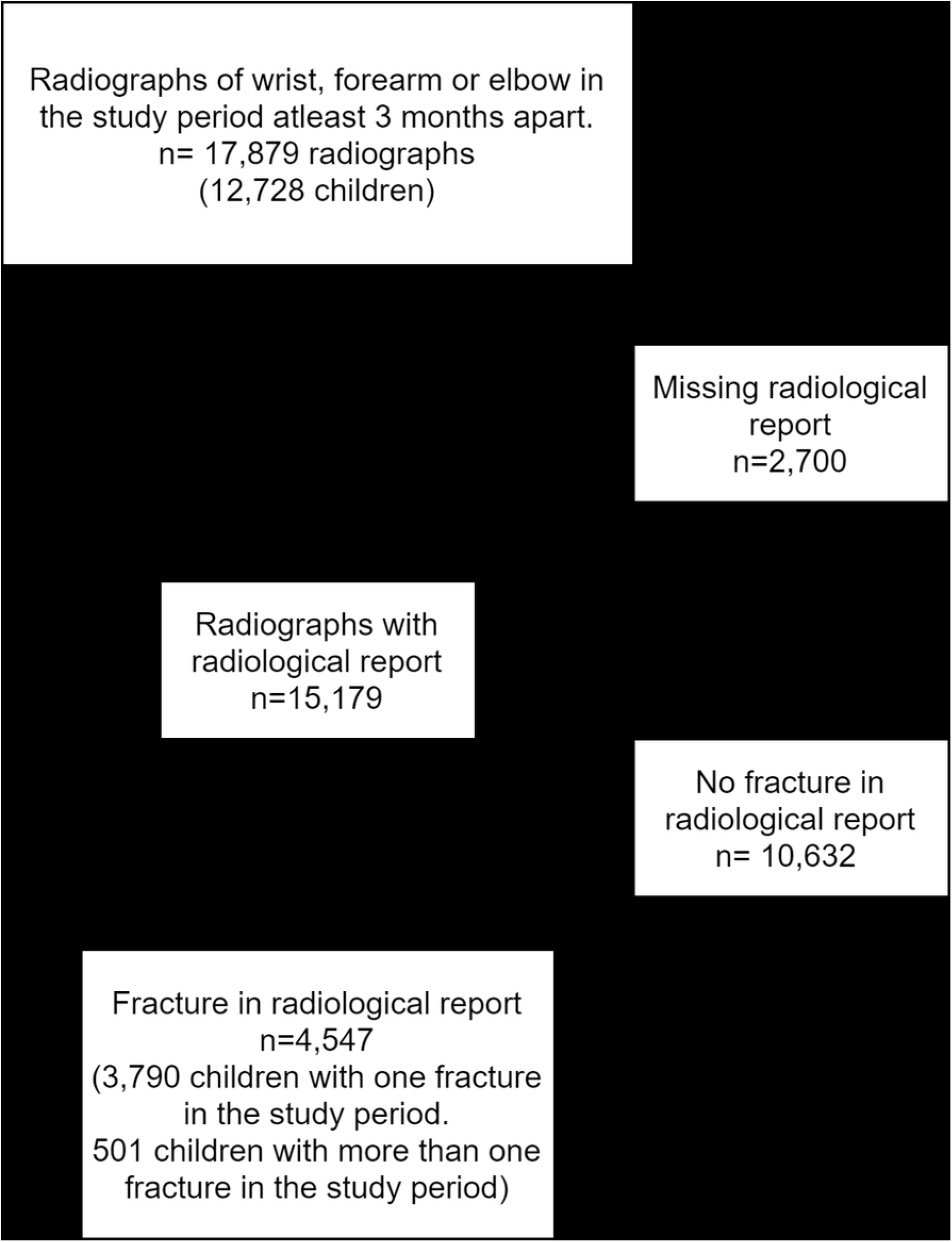
Stacked barchart of fracture location, fractured bone, fracture type and epiphyseal fracture status stratified by age group. The number of fractures within each age group were 0-3 years: n=335, 4-7 years: n=1,143, 8-11 years: n=1,679, 12-15 years: n=1,390.

For younger children, we observed proportionally fewer isolated radial fractures, distal fractures, complete fractures, and epiphyseal fractures (Table 1, Figure 2). Distribution of SH types did not differ across age groups (p=0.69).

## Discussion

This study provides a comprehensive overview of paediatric forearm fracture patterns based on radiological reports over a five-year period, finding that fracture patterns of the forearm in children vary significantly between age groups with younger children having fewer complete fractures, isolated radial fractures, distal fractures, and epiphyseal fractures.

### Interpretation

With increasing age, the fractures in the study population became increasingly concentrated in the distal forearm, with a higher proportion of complete fractures, radial involvement, and epiphyseal fractures. This shift likely reflects the changes in bone composition and biomechanics during growth ^13^, as well as activity patterns, which change as children move through different developmental stages ^14,15^. Younger children, who experience more incomplete fractures and fractures further from the epiphysis, may be more prone to these due to the pliability of their bones and reduced exposure to high-impact trauma compared to adolescents.

This dominance of distal forearm fractures aligns with the existing literature, which emphasizes the vulnerability of the distal radius and ulna, particularly in older children and adolescents ^4,16^. Our study found that adolescent boys were more frequently affected, consistent with previous studies that highlight the role of physical activity and risk-taking behaviour in this group ^1,4,15^.

While this study found that the prevalence of epiphyseal fractures increases with age, no significant difference in the distribution of Salter-Harris (SH) classifications across age groups was found. There was, however, a considerable proportion (>10%) of undescribed epiphyseal fractures for children above the age of eight years. This underreporting, combined with the relatively low number of physeal fractures, might be the cause of our inability to detect a significant difference in SH classification between age groups and might represent a type-2 error, rather than a true difference in age-specific prevalence in the background population.

### Strengths and limitations

While many of the findings of this study has been confirmed by previous studies, this is the first study, to our knowledge, to give a comprehensive accessible overview of fracture patterns of the entire forearm in a paediatric study population and might serve as a useful reference benchmark for future studies.

Methodologically, the study’s strength lies in its reliance on radiological reports, providing a high level of detail compared to studies using broader diagnostic codes such as ICD-10. The manual extraction of data allowed for precise categorization of fracture types and locations, which may be underreported in large-scale epidemiological databases. The exclusion of 15% missing radiological reports might introduce a selection bias, as radiographs without reports could represent less severe injuries, which might increase the calculated fracture proportions. Additionally, the exclusion of suspected fractures or uncertain cases and the three-month interval between fractures may have led to an underreporting of some types of paediatric forearm fractures.

Despite these limitations, this paper represents the largest study to date of manually extracted data of paediatric forearm fractures. By relying on written radiological reports, we were able to gather detailed information on fracture types and distributions.

### Generalizability

This study was conducted in a single region in Denmark, but as the distribution of fractures across age groups resemble those from national demographic studies ^9^, we believe the results to be valid for the entire population.

## Conclusion

This study offers a detailed characterization of paediatric forearm fractures, emphasizing the variability in fracture patterns with age. The findings underscore the importance of age-specific considerations in both clinical management and preventive strategies. Further research should build on these findings, exploring the implications of these fracture patterns on long-term functional outcomes and refining guidelines for the treatment and prevention of paediatric forearm fractures.

## Data Availability

Due to the sensitive nature of the data, it is not freely available. However, data can be shared upon reasonable request, subject to institutional approval and the signing of a data sharing agreement. Interested researchers may contact the corresponding author for further details.

## Author Contributions

The authors contributed to the research and manuscript preparation as follows:

- Study Conceptualization and Design: HCH, SK, OR
- Data Collection: HCH
- Data Analysis and Interpretation: HCH, SK, OR
- Manuscript Drafting and Writing: HCH, SK, OR
- Critical Review and Revisions: SK, OR
- Resources and Funding Acquisition: SK, OR

All authors read and approved the final manuscript and agree to be accountable for the work.

## Ethical Statement

This study was reviewed by the North Denmark Region Ethical Committee, which determined that ethical approval was not required (no: 2-1-02-1167-24).

